# Real-life protection provided by vaccination, booster doses and previous infection against covid-19 infection, hospitalisation or death over time in the Czech Republic: a whole country retrospective view

**DOI:** 10.1101/2021.12.10.21267590

**Authors:** Luděk Berec, Martin Šmíd, Lenka Přibylová, Ondřej Májek, Tomáš Pavlík, Jiří Jarkovský, Milan Zajíček, Jakub Weiner, Tamara Barusová, Jan Trnka

**Author notes:** Correspondence to: Luděk Berec, Centre for Mathematical Biology, Institute of Mathematics, Faculty of Science, University of South Bohemia, Branišovská 1760, 37005 České Budějovice, Czech Republic.

## Abstract

**Background:** Evidence is accumulating that the effectiveness of covid-19 vaccines against infection wanes, reaching relatively low values after 6 months. Published studies demonstrating this effect based their findings on a limited range of vaccines or subset of populations, and did not include booster vaccine doses or immunity obtained due to covid-19 infection. Here we evaluate effectiveness of covid-19 vaccines, booster doses or previous infection against covid-19 infection, hospital admission or death for the whole population in the Czech Republic.

**Methods:** Data used in this study cover the whole population of the Czech Republic reported as infected and/or vaccinated between the first detected case on March 1, 2020 and November 20, 2021 (for reinfections), or December 26, 2020 and November 20, 2021 (for vaccinations), including hospital admissions and deaths. Vaccinations by all vaccines approved in the EU were included in this study. Anonymous, individual-level data including dates of vaccination, infection, hospital admission and death were provided by the the Institute of Health Information and Statistics of the Czech Republic. The risks of reinfection, breakthrough infection after vaccination, hospital admission and death were calculated using hazard ratios from a Cox regression adjusted for sex, age, vaccine type and vaccination status.

**Findings:** The vaccine effectiveness against any PCR-confirmed infection declined from 87% (95% CI 86-87) at 0-2 months after the second dose to 53% (95% CI 52-54) at 7-8 months for Comirnaty, from 90% (95% CI 89-91) at 0-2 monthsto 65% (95% CI 63-67) at 7-8 months for Spikevax, and from 83% (95% CI 80-85) at 0-2 months to 55% at (95% CI 54-56) 5-6 months for the Vaxzevria. For Janssen Covid-19 Vaccine we found no significant decline but the estimates are less certain. The vaccine effectiveness against hospital admissions and deaths decayed at a significantly lower rate with about 15%, resp. 10% decline during the first 6-8 months. The administration of a booster dose returns the protection to or above the estimates in the first two months after dose 2. In unvaccinated but previously SARS-CoV-2-positive individuals the protection against PCR-confirmed SARS-CoV-2 infection declined from close to 97% (95% CI 97-97) after 2 months through 90% at 6 months down to 72% (95% CI 65-78) at 18 months.

**Interpretation:** Our results confirm the waning of vaccination-induced immunity against infection and a smaller decline in the protection against hospital admission and death. A booster dose is shown to restore the vaccine effectiveness back to the levels seen soon after the completion of the basic vaccination schedule. The post-infection immunity decreases over time, too.

**Funding:** No external funding was used to conduct this study.

**Research in context:** *Evidence before this study:* Accumulating evidence from several countries indicates that the effectiveness of covid-19 vaccines against infection declines in time, from about 80-90% shortly after completing the vaccination to about 50-60% and even less after 6 months. Published studies also suggest a significant boosting in vaccine effectiveness against infection about one week after the third vaccine dose. However, these observations come from different and often limited data sets. Moreover, the existing studies do not compare the decline in vaccine effectiveness with a decline in infection-based immunity in unvaccinated individuals.

*Added value of this study:* In our study, we bring together data on infections, vaccinations (including booster doses), hospital admissions and deaths to estimate how the protection due to vaccination or previous SARS-CoV-2 infection declines with time, for the whole population of the Czech Republic. Our findings show an overall decrease in vaccine effectiveness over time and a large increase after the administration of a booster dose. At the same time we show a fairly stable and high post-infection immunity over the study period. We hope this evidence will contribute to a better understanding of the changing impact of vaccines and previous infection in complex, real-world environments, which is crucial for the development of more effective and more easily communicated public health policies.

*Implications of all the available evidence:* Our results strongly support a timely and widespread application of booster vaccine doses since their application appears to restore the vaccine-induced protection to the levels attained soon after completing the original vaccination scheme, including the high protection against mild disease or asymptomatic infection.

## 1. Introduction

The availability of vaccines brought about a breakthrough in the fight against the coronavirus disease 2019 (covid-19) worldwide. In the light of the economic and social costs already caused by covid-19, and the widespread aversion to any serious limitations of people’s daily lives due to lockdowns, vaccination is undoubtedly a key tool for the containment of the pandemic and for the limiting of its devastating impact on lives and health of people around the globe. Proving in the clinical studies and the weeks and months of their real-world application their high effectiveness against SARS-CoV-2 infection, symptomatic covid-19 illness, need of a hospital admission, the probability of severe symptoms, and death (Polack *and others*, 2020; Baden *and others*, 2021; Voysey *and others*, 2021; Sadoff *and others*, 2021), their continued impact now starts to be challenged by an increasing proportion of breakthrough infections and illnesses in fully vaccinated individuals (Brown *and others*, 2021; Levine-Tiefenbrun *and others*, 2021) and the appearance of new waves of infection in many world regions.

In the Czech Republic, vaccination started on December 27, 2020 initially with the mRNA-based vaccine Comirnaty (BNT162b2, Pfizer/BioNTech), followed by Spikevax (mRNA-1273, Moderna), and the adenovirus-based vector vaccines Vaxzevria (ChAdOx1 nCoV-19, AstraZeneca) and Janssen Covid-19 Vaccine (Ad26.CoV2.S, Johnson&Johnson, further abbreviated as Janssen). The rate of vaccination was increasing until the beginning of June, 2021, stayed relatively low during summer months, and started to rise again in October 2021, in response to epidemic control measures requiring a proof of infection/vaccination/negative test in many public places (Ministry of Health of the Czech Republic, 2020). The administration of booster doses in the Czech Republic started on September 20, 2021 and was initially open to all individuals who completed their vaccination 8 months or longer ago with only Comirnaty and Spikevax as allowed boosting vaccines. The waiting period was shortened to 6 months on October 29, 2021. Until November 1, 2021 a full 100 µg dose of Moderna was being administered as a booster dose and since that day only a 50 µg dose was used.

Similar to many other world regions, Central Europe experienced another wave of SARS-CoV-2 infections in the autumn of 2021 despite the substantial proportion of vaccinated and/or recovered population. This wave was accompanied by a non-negligible proportion of new infection cases in vaccinated individuals including the need for hospital admission and, in a relatively few cases, for intensive care. The appearance of breakthrough infections, though not unexpected, has complicated the public health messaging related to the importance of vaccination and calls for a better understanding of the temporal dynamics of post-vaccination immunity in real-world settings.

Post-infection immunity is another factor, which plays an important role in the population dynamics of viral transmission and is an important determinant of individual risk. SARS-CoV-2 reinfections have been reported as relatively rare events, yet the post-infection immunity appears to wane, too (Abu-Raddad and Bertollini, 2021; Cavanaugh *and others*, 2021; Hansen *and others*, 2021; Townsend *and others*, 2021). However, none of these studies addressed longer-term dynamics of post-infection immunity and their relationship to post-vaccination immunity.

In this study, we set out to estimate the extent of the waning of post-vaccination and postinfection immunity against SARS-CoV-2 infections, covid-19 hospital admissions, and deaths in the whole population of the Czech Republic and assess the influence of the type of vaccine and previous PCR-confirmed SARS-CoV-2 infection.

## 2. Methods

### 2.1 Study population and data sources

The analyses are based on data from the Czech National Information System of Infectious Diseases (ISID), which includes records of all individuals tested positive for SARS-CoV-2 in the Czech Republic since the beginning of covid-19 pandemic, including children (Komenda *and others*, 2020). This database is overseen by the Czech Ministry of Health and operated by the Institute of Health Information and Statistics of the Czech Republic. The ISID data is routinely collected in compliance with the Act No. 258/2000 Coll. on the Protection of Public Health. The data collection was managed and their usage approved by the Institute of Health Information and Statistics of the Czech Republic, according to the rules set by the Ministry of Health of the Czech Republic. According to a decision of the Institute of Health Information and Statistics of the Czech Republic, our retrospective analyses did not require ethical approval. Among other things, the ISID database covers demographic data, dates of vaccination, including the vaccine types for each dose, and dates of infection and potential reinfection, including information on dates of hospital admission with covid-19, and death with covid-19. Additional information on deaths from any cause come from the Death Certificate System; these data are used for censoring purposes only.

In total, our dataset contains 7,428,968 valid records of vaccinated and/or SARS-CoV-2 positive persons (additional 8,834 cases lack information on sex or age and 216 other cases contain data errors, see Table S4). We further excluded 16,399 persons who were recorded to die by the start of vaccination (December 26, 2020). As the source dataset consists only of those who were tested positive and/or were vaccinated, we completed the sample to the whole population such that the added subjects were neither tested positive nor vaccinated. In particular, we completed each sex-age category to the numbers reported by the Czech Statistical Office by December 31, 2020 – 10,701,777 inhabitants; consequently, our sample truly reflected the sex and age structure of the whole population, containing all the positive and/or vaccinated individuals. We neglected births and deaths of the added persons.

### 2.2 Vaccine types and vaccination and infection dynamics

In the Czech Republic, all EMA-approved Covid-19 vaccines have been distributed and used. They were provided to all individuals at no cost following the Czech public health insurance system. Starting on December 27, 2020, workers in the critical infrastructure were vaccinated first, followed since January 15, 2021 by persons of age 80 and older (Table S3 in the Supplementary material). As of November 20, 2021, the national Institute of Health Information and Statistics reported 6,287,356 individuals completing the vaccination (58.75% of the population and 67.36% of persons of age 12 years and older); see Fig. 1.

**Fig. 1.**
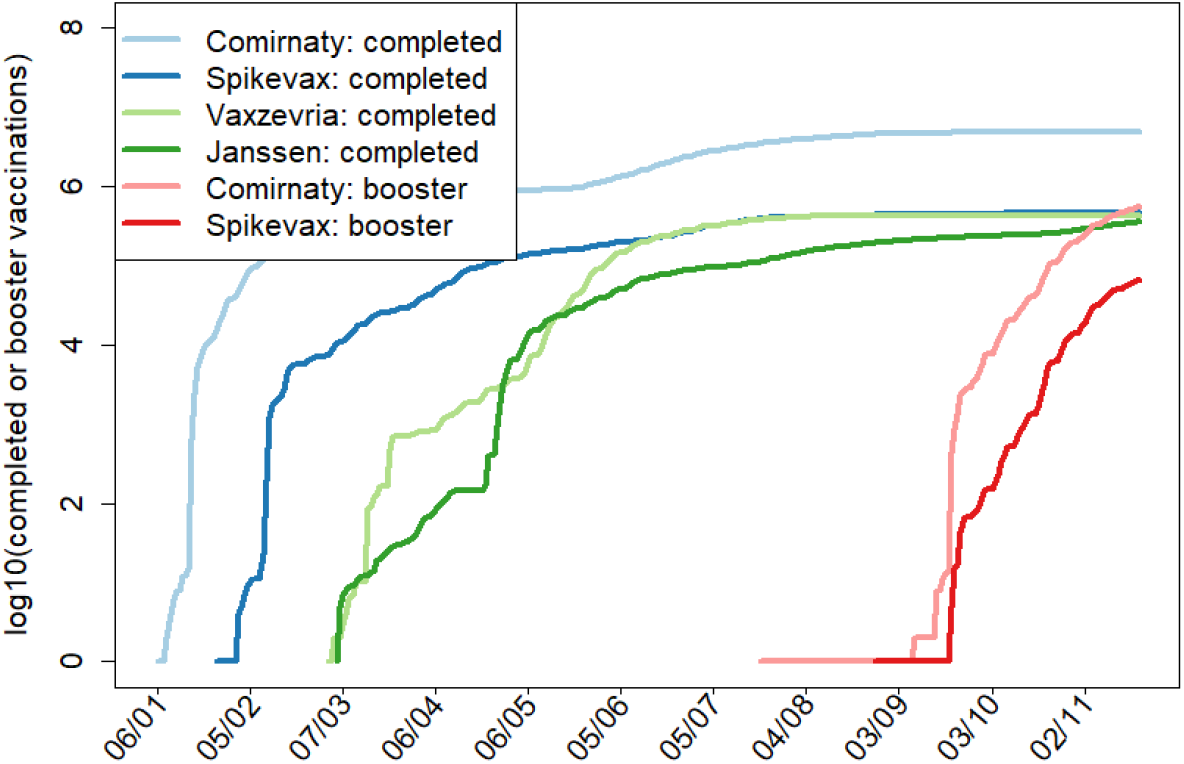
Dynamics of vaccination in the Czech Republic. Specific days in which vaccination was open to an age group or professional or other category are specified in Table S3 in the Supplementary Material.

The covid-19 epidemic in the Czech Republic started with the first three cases reported on March 1, 2020 and was initially fueled by Czech citizens returning from the alpine ski resorts of Italy and Austria. Since then the country saw five waves of covid-19 spread. As of November 20, 2021, 1,996,080 individuals were infected with SARS-CoV-2 virus, of which 12,894 (0.65%) were reinfected; see Fig. S1. See also Tables S4 and S5 for an overview of the numbers of infectionrelated outcomes and vaccines applied within various age cohorts.

### 2.3 Statistical analysis

We separately studied three types of events: (i) SARS-CoV-2 infection defined as a PCR confirmed positive test of a person from any sample regardless of the presence of symptoms, (ii) hospital admission of a person tested positive via a PCR test (within two weeks before hospital admission and whenever during hospitalization), and (iii) death due to covid-19.

A Cox regression with time-varying covariates was applied to estimate hazard ratios (HRs) for the outcomes of interest. Analogously to Tartof *and others* (2021), we used calendar time instead of time from event occurrence as the time scale. Thus, the time course of individual cases was modelled by means of “switching” dummy variables, corresponding to the stages of the process the subject goes through. The vaccine effectiveness is calculated by comparing hazards of the vaccinated individuals to those of the control group – those who have not been vaccinated and infected so far – individually for each vaccine type, following Tartof *and others* (2021). By using calendar time, we could automatically control for changing conditions of the epidemics, including adopted non-pharmaceutical measures, seasonal effects and virus variants, as all these phenomena can then be encompassed in the baseline hazard function.

In particular, time zero corresponded to the day before the start of vaccination (i.e., 26 December 2020) for the analyses of vaccine-induced immunity, and the onset of epidemic in the Czech Republic for the analyses of infection-induced immunity. Moreover, we estimated how HRs of infection after vaccination depended on time after the vaccine application (adjusted for sex, age and time since the last infection), how HRs of hospital admission or death depended on time after the vaccine application (adjusted for sex and age), and how HRs of reinfection in unvaccinated individiuals depended on time since the previous infection (adjusted for sex and age). In all these cases, we estimated the vaccine effectiveness (VE, regarding a previous infection as a “vaccine”) as VE = 1 − HR (Chemaitelly *and others*, 2021; Cohn *and others*, 2021; Tartof *and others*, 2021). Subjects were withdrawn from the study at the time of their (covid or non-covid) death.

We aggregate the time delays in two-month (61 days) periods. In particular, we consider one such period after the first dose, four periods after the second one and a single period after the booster dose. When a new dose is applied to a person, (s)he is no longer regarded to be in any period corresponding to the previous dose, but enters the first period corresponding to the new dose. In line with the Czech vaccination recognition policy, the first period corresponding to any of the first two doses starts two weeks after the dose application, while for boosters this interval is just 7 days. For the reinfections, we consider nine two-month periods.

Since there are only several hundreds of booster doses of Vaxzevria and Janssen vaccines, we examine boosting effects by the mRNA vaccines Comirnaty and Spikevax only. Since 93% of Comirnaty boosters is preceded by the Comirnaty second dose and 73% of Spikevax boosters is preceded by the Spikevax second dose, but the vaccine type used for the first and the second doses does not play a role in which vaccine type is applied as the booster, we estimate the boosting effects of the mRNA vaccines both with and without individual vaccination history. Subjects with the alleged application of Vaxzevria (in total 176) or Janssen (in total 332) boosters are thus withdrawn from the study of booster dose effectiveness, as these records are most likely data entry errors.

To analyse a possible impact of the delta variant to breakthrough infections, we made an alternative analysis including dummy variables indicating time period starting on July 1st, 2021 when the delta variant started to dominate in the Czech Republic (virus.img.cas.cz/lineages). We performed three such comparisons, each concerning only age cohorts which started to be extensively vaccinated at similar time (see Table S3); only this way we could guarantee to some extent that the estimation of the delta effect would not collide with that of immunity waning.

All calculations were performed using the R software (package `survival`). The algorithm used to transform data from the database into the package command inputs was coded in C++. See Supplementary material for details.

## 3. Results

Since December 26, 2020 (one day before start of vaccination) to November 20, 2021, 6,287,356 individuals received full vaccination (58.75% of the population and 67.36% of persons of age 12 years and older). In this period a total of 1,335,055 individuals were infected, of which 96,237 (7.21%) were hospitalized and 20,809 (1.56%) died because of covid-19 (Table S4). Out of the vaccinated individuals by far the largest group of 5,011,115 persons (79.7%) received Comirnaty, followed by similarly sized groups of 469,605 persons (7.47%) vaccinated with Spikevax, 436,575 persons (6.94%) with Vaxzevria and 370,061 persons (5.89%) with the one-dose Janssen vaccine (Table S5). The 693,071 booster doses administered in this period comprised 617,002 doses of Comirnaty and 76,069 doses of Spikevax (see Table S5 for details).

Using an age-adjusted Cox model we estimated the change in vaccine effectiveness over time at two-month intervals (Fig. 2, Table S6 in Supplementary Material). The vaccine effectiveness against any PCR-confirmed SARS-CoV-2 infection declined for Comirnaty from 87% (95% CI 86-87) 0-2 months after the second dose to 53% (95% CI 52-54) at 7-8 months, for Spikevax from 90% (95% CI 89-91) at 0-2 months to 65% (95% CI 63-67) at 7-8 months, and for Vaxzevria from 83% (95% CI 80-85) at 0-2 months to 55% (95% CI 54-56) at 5-6 months. Interestingly, the estimated effectiveness for the Janssen vaccine (68% (95% CI 66-70) at 0-2 months and 67% (95% CI 65-69) at 5-6 months) did not seem to exhibit any significant decline over the study period but notably starts at a significantly lower effectiveness (Fig. 2 blue curves, Table S6). The effectiveness estimates for Vaxzevria and Janssen at 7-8 months after the completion of vaccination exhibit very large uncertainty due to a low number of events as most people completed their vaccination with these vaccines much later, and are therefore only shown in Table S6.

**Fig. 2.**
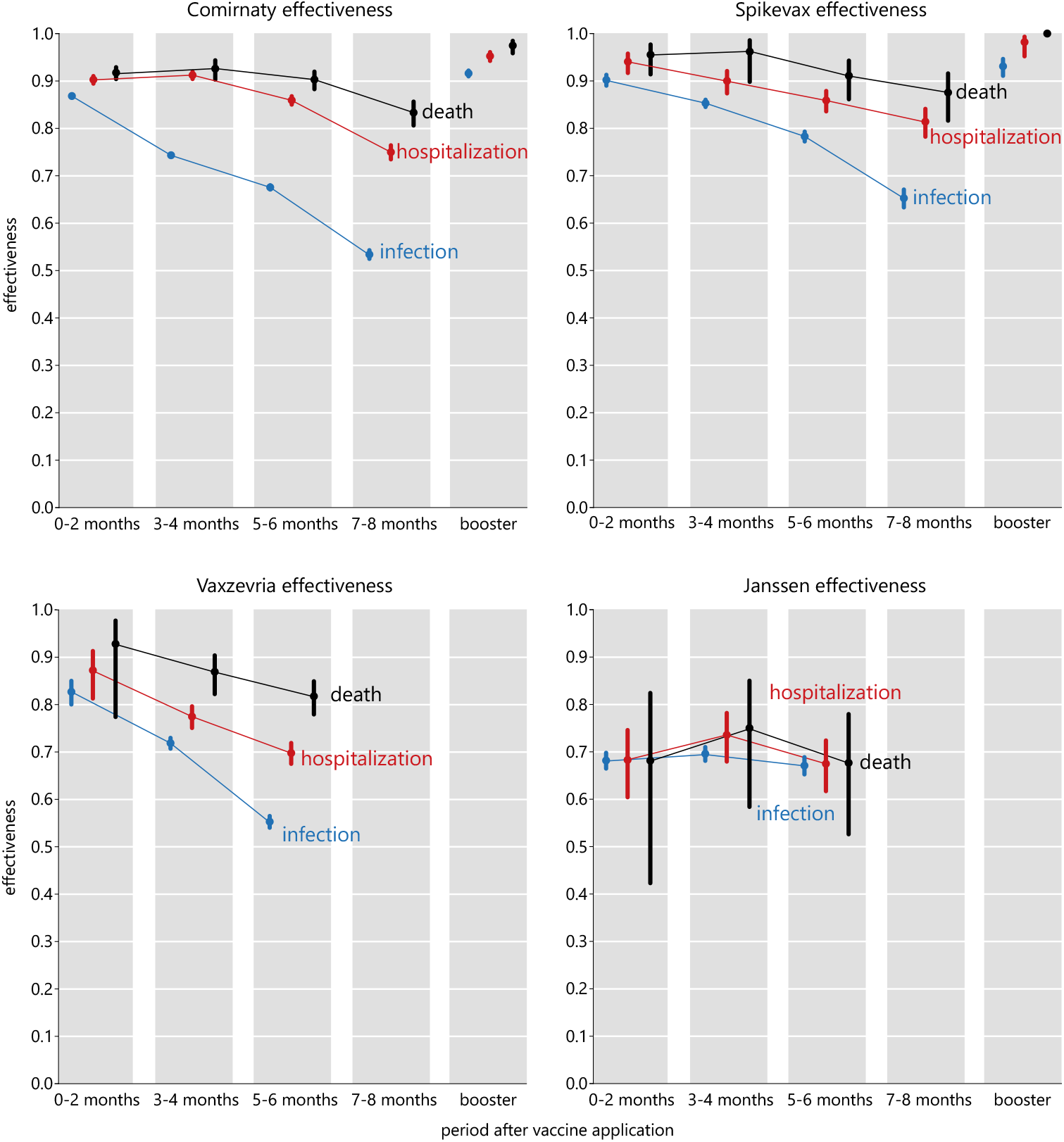
Vaccine-acquired immunity against infection with respect to the delay from the full vaccine application, including the effect of a booster vaccine dose.

A similar trend can be seen in the estimation of vaccine effectiveness against hospital admissions and deaths. For hospital admission, the vaccine effectiveness declined for Comirnaty from 90% (95% CI 89-91) at 0-2 months after dose 2 to 75% (95% CI 73-76) at 7-8 months, for Spikevax from 94% (95% CI 92-96) to 81% (95% CI 78-84), and for Vaxzevria from 87% (95% CI 81-91) at 0-2 months to 70% (95% CI 68-72) at 5-6 months (Fig. 2 red curves, Table S6). In the case of protection from death the model estimated for Comirnaty a decrease from 92% (95% CI 90-93) at 0-2 months to 83% (95% CI 81-86) at 7-8 months, from 96% (95% CI 91-98) to 88% (95% CI 82-92) for Spikevax within the first 8 months and from 93% (95% CI 77-98) to 82% (95% CI 78-85) for Vaxzevria within the first 6 months after application (Fig. 2 black curves, Table S6). Janssen once again exhibits virtually no decline either in the protection against hospitalization starting from 68% (95% CI 60-75) at 2 months to 67% (95% CI 62-72) at 5-6 months, or deaths starting from 68% (95% CI 42-82) and reaching 68% (95% CI 53-78) at 5-6 months (Fig. 2, Table S6).

In June-July 2021 the alpha variant of the SARS-CoV-2 virus was largely superseded by the delta variant in the Czech Republic (virus.img.cas.cz/lineages). Therefore, we attempted to disentangle the effects of immunity waning and immunity evasion due to the delta variant on the observed changes in the vaccine effectiveness. Evaluating the extra risk of breakthrough infection due to the delta variant, for consistency estimated just for the age cohorts that started to be vaccinated at about the same time, we found a consistent and significant increase in the risk for Comirnaty, mostly significant increase for Spikevax and Vaxzervia, and inconclusive results for Janssen (Table 1). Note that these differences do not represent the infection risk increase due to delta; they represent the additional risk increase of a vaccinated individual over the generally higher infectiousness of the delta variant compared to alpha.

**Table 1.**
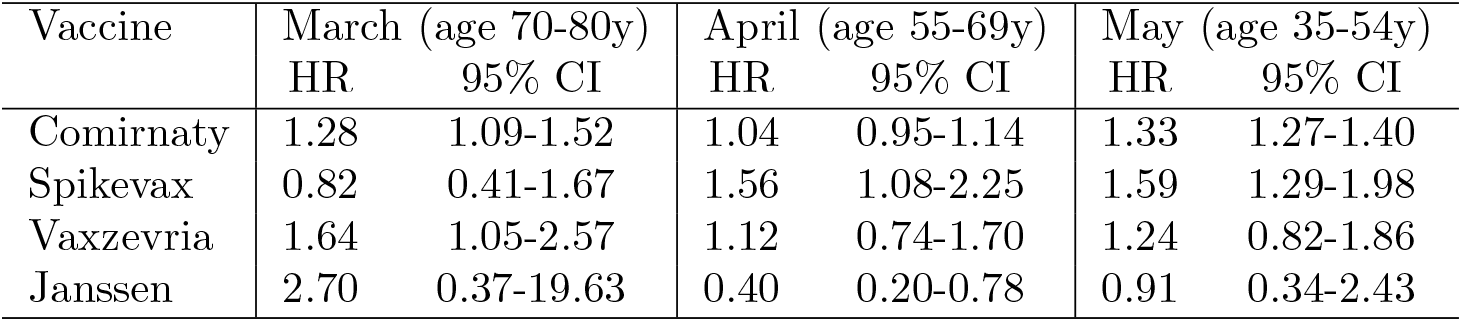
Estimated increase of breakthrough infection hazard ratios (HRs) in times of the SARS-CoV-2 delta variant dominance for age groups having started vaccination in the same month.

| Vaccine | March (age 70-80y) |  | April (age 55-69y) |  | May (age 35-54y) |  |
| --- | --- | --- | --- | --- | --- | --- |
|  | HR | 95% CI | HR | 95% CI | HR | 95% CI |
| Comirnaty | 1.28 | 1.09-1.52 | 1.04 | 0.95-1.14 | 1.33 | 1.27-1.40 |
| Spikevax | 0.82 | 0.41-1.67 | 1.56 | 1.08-2.25 | 1.59 | 1.29-1.98 |
| Vaxzevria | 1.64 | 1.05-2.57 | 1.12 | 0.74-1.70 | 1.24 | 0.82-1.86 |
| Janssen | 2.70 | 0.37-19.63 | 0.40 | 0.20-0.78 | 0.91 | 0.34-2.43 |

Regardless of the original vaccine used for the initial vaccination schedule a Comirnaty booster dose brings the protection against infection to 92% (95% CI 91-92), against hospital admission to 95% (95% CI 94-96), and against death to 97% (95% CI 96-98) (Figs 2). A Spikevax booster dose reaches 93% (95% CI 91-95) protection against infection, 98% (95% CI 95-99) against hospital admission, and close to 100% against death (Figs 2). When we looked at the various possible combinations of primary and booster vaccines (with the exception of Janssen due to insufficient data) we found that a combination of two mRNA-based vaccines reached a boosted effectiveness of *>* 91%. The combination of Vaxzevria primary and an mRNA booster showed a somewhat lower effectiveness but these estimates are less certain due to a low number of observations (Table 2).

**Table 2.**
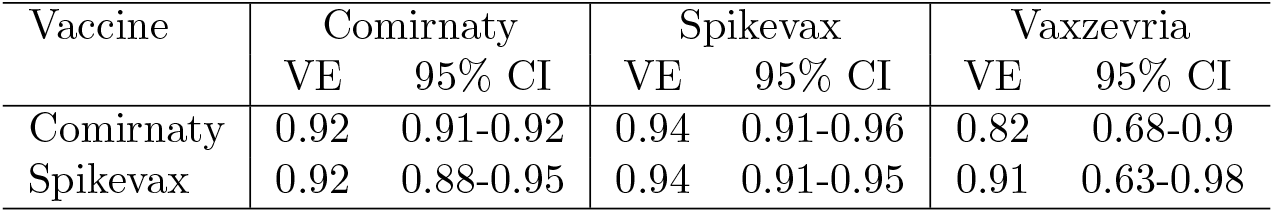
Vaccine effectiveness against infection after administering the booster vaccine dose for various possible combinations of primary (columns) and booster (rows) vaccines (with the exception of Janssen due to insufficient data). Hazard ratios (HRs) are given.

| Vaccine | Comirnaty |  | Spikevax |  | Vaxzevria |  |
| --- | --- | --- | --- | --- | --- | --- |
|  | VE | 95% CI | VE | 95% CI | VE | 95% CI |
| Comirnaty | 0.92 | 0.91-0.92 | 0.94 | 0.91-0.96 | 0.82 | 0.68-0.9 |
| Spikevax | 0.92 | 0.88-0.95 | 0.94 | 0.91-0.95 | 0.91 | 0.63-0.98 |

For the study of reinfections we used data on PCR-confirmed infections since the beginning of covid-19 pandemic in the Czech Republic; 1,999,315 individuals were infected with SARS-CoV-2 virus until 20th November 2021, of which 12,894 (0.64%) were reinfected. Previous SARS-CoV-2 infection in our population conferred a high and fairly stable level of protection against infection lasting for more than 18 months. In unvaccinated but previously covid-19-positive individuals the protection against PCR-confirmed covid-19 infection declined from close to 97% (95% CI 97-97) at 2-4 months through 91% (95% CI 90-91) at 5-6 months down to 83% (95% CI 82-84) at 11-12 months and 72% (95% CI 65-78) at 17-18 months (Fig. 3, Table S7).

**Fig. 3.**
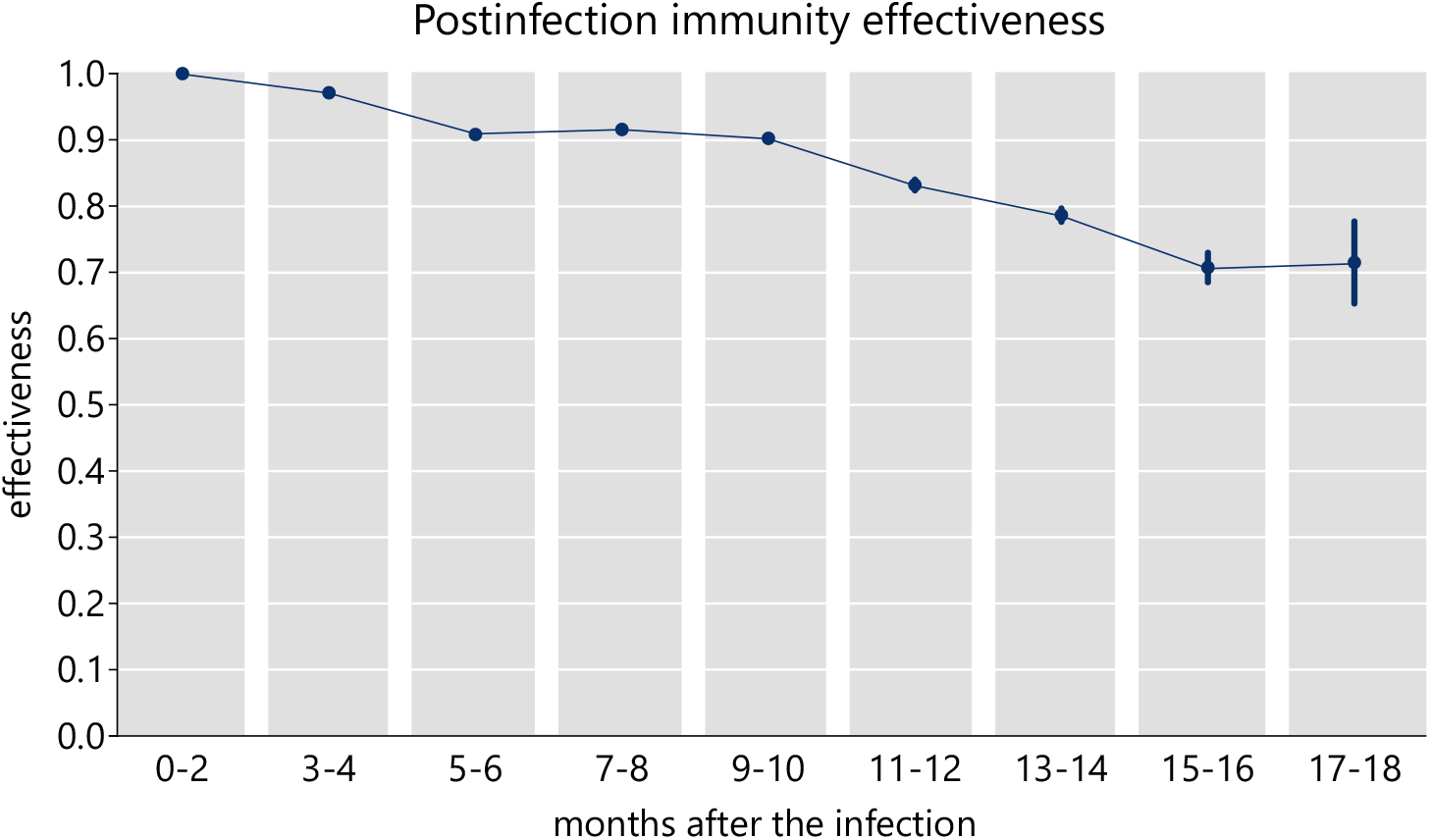
Infection-acquired immunity against reinfection with respect to the delay from the prior infection. The delay 0-2 months is not considered as a new infection which implies 100% effectiveness by definition.

## 4. Discussion and Conclusions

Results presented here show a gradual decrease in the protective effectiveness of three out of the four vaccines used in the Czech Republic to vaccinate against covid-19. The observed decrease in the protective effect of vaccines was the fastest for the protection against infections followed by hospital admissions, while the protection from covid-19 related death was the least affected by the time elapsed from the completion of the primary vaccination schedule.

There are several plausible explanations for this decrease and for a corresponding rise in breakthrough infections. One of them is a waning of the immunity conferred by the vaccines, which has been documented for a range of commonly used vaccines and has been demonstrated for covid-19 vaccines in an increasing number of recent studies from several countries (Chemaitelly *and others*, 2021; Cohn *and others*, 2021; Tartof *and others*, 2021). The other possible contributor is the effect of the delta variant, which has been shown to evade to some extent the vaccine-induced immunity (Lopez Bernal *and others*, 2021; Tang *and others*, 2021; Tartof *and others*, 2021). Our sub-analysis of the data suggests but a modest overall effect of the delta variant on the vaccine effectiveness in the studied period. Not having individual-level information about the specific variant causing a breakthrough infections we used in this sub-analysis an indirect method of time dummies corresponding to the period of delta variant dominance. We are aware that this approach may be affected by co-linearity (namely that the “influence” of the absolute time will be mismatched with the waning) and therefore we present these estimates only as secondary results and the primary effectiveness estimates are averaged over the viral variants. However, it is likely that long-term estimates of vaccine effectiveness correspond to the period of dominating delta variant. This is not an issue for boosters, which have been given only after delta reached overwhelming dominance.

A further and largely unstudied factor that could affect the observed vaccine effectiveness are changes in the behaviour of vaccinated compared to unvaccinated persons and the possible effect of infection control measures due to a differential access of the unvaccinated individuals to many social activities or due to differential testing strategies, through which vaccinated people and people within 6 months after their PCR-positivity have not been required to undergo testing as often as the others. Indeed, analyses of vaccine effectiveness and its temporal dynamics generally assume that both vaccinated and unvaccinated persons behave similarly and we assume this in our study as well. This is a possible limitation of this study, applicable mainly to the endpoint of confirmed infection and to the lesser extent to the hospitalisation or death endpoints.

The one-dose Janssen vaccine appears in our analysis to defy the general trend of protection decay and while it starts at a significantly lower effectiveness, it holds it over the six months analysed in this study. To our knowledge, this somewhat counter-intuitive result has not yet been reported and as such it is not easy to interpret. However, due to the fact that this vaccine was introduced to the Czech Republic much later than the other three vaccines and that it requires only one dose for a complete vaccination, it is plausible to assume that this vaccine was mostly chosen by people with different social and behavioural characteristics compared to the two-dose vaccines. Since we cannot support this suggestion with data at the moment we leave this as a suggestion for further studies.

Our results show that the administration of booster doses of the two approved mRNA vaccines brings the observed effectiveness to above 90% for infections, hospital admissions and deaths alike. Booster doses are undoubtedly highly efficient to prevent serious or fatal infections. Although our results are in a general agreement with the study on protective effect of vaccine booster in Israel (Bar-On *and others*, 2021), we cover a more extensive period of booster applications, use of the Spikevax vaccine as a booster, and do not limit ourselves to a specific age group.

The protection afforded by a previous covid-19 infection declines over time too but at a significantly slower rate compared to the post-vaccination immunity. Whereas several studies consistently report that protection against reinfection declines (Abu-Raddad and Bertollini, 2021; Hansen *and others*, 2021; Cavanaugh *and others*, 2021; Townsend *and others*, 2021), we are the first to describe the long-term temporal dynamics of infection-induced immunity against SARS-CoV-2 reinfections. However, especially in the light of the currently spreading omicron variant of the SARS-CoV-2 virus we warn against any simple conversion of the observed reinfection risks into public health policies. It is also necessary to acknowledge that this finding relates only to directly confirmed primary infections (possibly associated with test-seeking behaviour and severity of the disease) and may not be translatable to evidence of previous infection from antibody testing.

In this study we used a Cox model with calendar time, which has an obvious advantage – our results are independent of factors that influence the risk of all the subjects equally, such as a change in the basic reproduction number, viral prevalence in the population, non-pharmaceutical interventions, weather or seasonal influences, or a dominant virus variant – all these factors can be included in the baseline hazard function of the Cox model. All this makes our findings comparable with and transferable to other contexts. Indeed, similar studies have come to similar conclusions Tartof *and others* (2021). Our results are also robust with respect to under-reporting provided the reporting rate is the same for all subjects, because then the Cox regression equation is only multiplied by a constant, so the estimation of the HRs remains correct.

The model, however, has some limitations. Importantly, the dependence of individual hazard function on covariates may be non-log-linear. This happens, for instance, when the detection rate depends on a characteristic of a subject – e.g. unvaccinated are being tested more often than the vaccinated. If this were true the vaccine effectiveness would be overestimated, yet the estimates of the HR increase (i.e. VE decline) over time would still be valid (provided that the testing propensity does not change in time). Equally such a case could arise if the vaccinated behaved more riskily than the unvaccinated – the effectiveness then would be underestimated, yet the estimates of the relative increase of HR (and the consequent decrease of VE) would again be valid provided that this behaviour does not change substantially over the study period. It is also worth noting that unlike infections the hospitalization and mortality data are less likely to suffer from the aforementioned bias as the facts of hospitalizations and deaths depend much less on test seeking behavior.

We also estimated the waning functions as constant over two-month intervals. We believe this approach is more valid than assuming a particular shape of the waning functions, for which we do not have a theoretical justification in the case of a relatively unexplored disease such as covid-19. When interpreting our results, however, it is necessary to bear in mind that not all subject pass through some time intervals so the resulting estimates of vaccine effectiveness are related the beginning rather than being an average over the whole duration. Consequently, the VE estimates will be higher than if these were computed as averages over the whole periods. In particular, this is the case for the period after the first dose of mRNA vaccines, the fourth period after the second doses and the boosters (see Table S8).

In summary, we used a comprehensive national population-based database containing individual level data about all detected SARS-CoV-2 infection cases to estimate many important characteristics of the post-vaccination and post-infection immunity in the population of the Czech Republic, covering all four vaccines currently approved in the EU and the protection from infection, hospital admission and death. The results strongly advocate for a timely and widespread administration of vaccine booster doses. Covid-19 will undoubtedly continue to disrupt everyday lives and cause suffering and loss of life around the globe and real-life vaccine effectiveness data such as the ones presented in this study can bring an important insight for policy makers in order to limit the worst impacts of the current pandemic.

## Data Availability

Data reported in this study and used for the analyses are not public. De-identified individual-level data are available to the scientific community. Requests should be submitted to the Institute of Health Information and Statistics of the Czech Republic (www.uzis.cz/index-en.php), together with a short description of their analysis proposals, where they will be assessed based on relevance and scientific merit.

## Contributors

LB, MŠ, and LP conceived this study. JJ, OM, JW, MZ, and TB prepared and checked the data, TP provided the methodological expertise, JW and MŠ conducted the analyses. LB and JT wrote the first draft of the manuscript. All authors contributed to writing the manuscript and interpreting the results. All authors approved the final version and had final responsibility for the decision to submit for publication.

## Supplementary material

### Supplementary figures

**Fig. S1.**
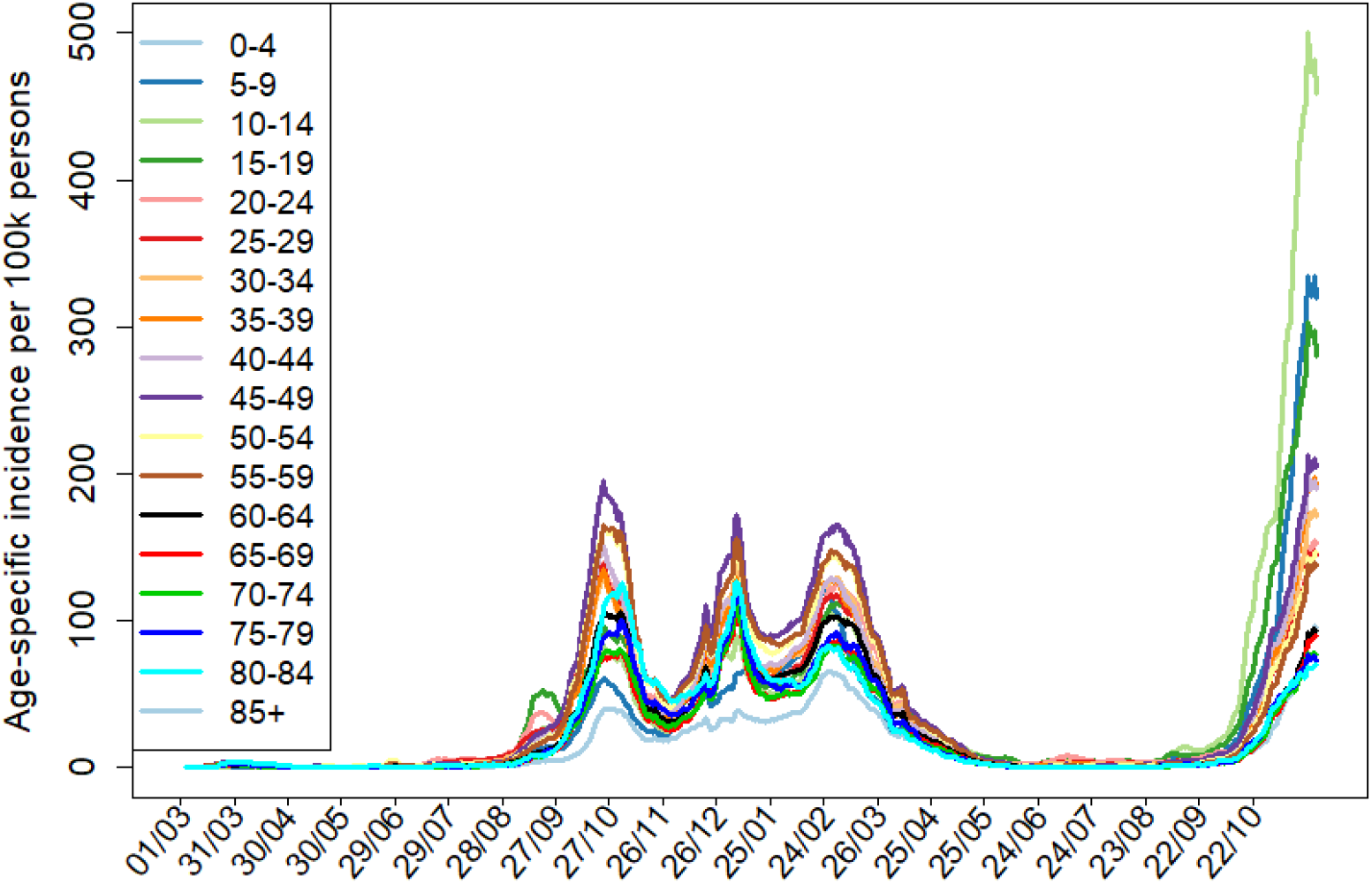
Daily incidence of covid-19 from March 1, 2020 till November 20, 2021 in the Czech Republic, stratified by age, per 100,000 individuals in the respective age groups.

### Methods: a Cox regression with time-varying covariates

We use a Cox proportional Hazard model with time varying covariates. In the model, we let each individual to go through the several “vaccination” (covariate `VaccStatus`) and “post-infection” (`InfPrior`) states. The outcomes (events) are either (a confirmed) infection (`Infected`, may be repeated), hospitalization (`Hospitalized`), or death of covid (`DeadByCov`). Deaths of other reasons are also recorded (`DeadByOther`), leading to withdrawal from the study at the time of the event. Fixed (non-time-dependent) covariates include sex (`Sex`) and age category (`AgeGr`). The input for the Cox regression model (`coxph` from `survival R` package) consists of one or more records for each subject, each referring to an interval from `T1` to `T2`, containing the values of covariates *Inf Prior, V accStatus, AgeGr, Sex* valid in [*T*1, *T*2) and indicators of outcomes `Infected, Hospitalized, DeadByCov, DeadByOther`, happening at *T* 2. There may be (and typically is) several records for each subject, each corresponding to a time interval in which the covariates are constant and in the interor of which no events happen.

Time is measured in days and we take the day before vaccination started (Dec 26th, 2021) as time zero in all analyses except for the reinfection analysis in which we take May 1st, 2020 (two months after the first cases) as time zero. The `VaccStatus` categorical covariate may take the following values:

`Unvacc`: The subject is not vaccinated (this value is taken as reference).

*V* `first1` The subject is from 14 to 14 + 61 − 1 = 74 days after the first dose of vaccine *V* but not 14 days or more after a second dose. *V* may be `A`–Vaxzevria, `M`–Spikevax or `P`–Comirnaty.

*V* `first2plus`: The subject is 75 days or more after the first dose of vaccine *V* but not 14 days or more after a second dose.

*VX* : The subject is between 14 + (*X* 1) 61 and 14 + (*X*) 61 1 days after the final dose of vaccine *V* but not 7 days or more after a booster. In addition to `P/M/A`, the vaccine may be also `J` (Janssen).

*V* `boost`: The subject is 7 days or more after a booster by vaccine *V*. The `InfPrior` may take the following values:

`None`: The subject has not been infected previously.

*X* : The subject is from (*X −* 1) × *p* to *X × p −* 1 days after the last positive test for covid, where *p* = 61 in the analyses of reinfections an *p* = 91 in the remaining analyses.

`rest`: In reinfection analysis: the subject is 9 × 61 = 549 days or more after the last positive test, in the remaining analyses: the subject is 3 × 91 = 273 days or more after the last positive test.

By default, for each subject, the intervals cover the entire period from 26th. December 2020 to 20. November 2021. The period is shortened if either

- The subject is reported to die

**Table S1.** Sample input to Cox regression.
- The subject is reported to obtain booster by Vaxzevria or Janssen
- The subject is 4*×*61+14 = 258 days after the final dose and has not yet obtained a booster.
- The subject is hospitalized (only in the hospitalization analysis)
- The subject is infected or gets a vaccine (only in the reinfection analysis)

| Sub-<br>ject | T1 | T2 | Inf-<br>ected | Dead-Covid | Dead-Other | Inf-Prior | Vacc-Status | Age-Gr | Sex |
| --- | --- | --- | --- | --- | --- | --- | --- | --- | --- |
| A0 | 0 | 313 | 1 | 0 | 0 | _none | _unvacc | 40-44 | F |
| A0 | 313 | 314 | 0 | 0 | 0 | 1 | _unvacc | 40-44 | F |
| A1 | 0 | 140 | 1 | 0 | 0 | _none | _unvacc | 75-79 | F |
| A1 | 140 | 150 | 0 | 1 | 0 | _none | _unvacc | 75-79 | F |
| A2 | 0 | 128(=114+14) | 0 | 0 | 0 | _none | _unvacc | 45-49 | M |
| A2 | 128 | 142 | 1 | 0 | 0 | _none | P_first1 | 45-49 | M |
| A2 | 142 | 189(=128+61) | 0 | 0 | 0 | 1 | P_first1 | 45-49 | M |
| A2 | 189 | 220 | 0 | 0 | 0 | 1 | P_first2plus | 45-49 | M |
| A2 | 220 | 233(=142+91) | 0 | 0 | 0 | 1 | P1 | 45-49 | M |
| A2 | 233 | 281(=220+61) | 0 | 0 | 0 | 2 | P1 | 45-49 | M |
| A2 | 281 | 314 | 0 | 0 | 0 | 2 | P2 | 45-49 | M |
| A3 | 0 | 71(=0-20+91) | 0 | 0 | 0 | 1 | _unvacc | 40-44 | F |
| A3 | 71 | 162(=71+91) | 0 | 0 | 0 | 2 | _unvacc | 40-44 | F |
| A3 | 162 | 164(=150+14) | 0 | 0 | 0 | 3 | _unvacc | 40-44 | F |
| A3 | 164 | 225(=164+61) | 0 | 0 | 0 | 3 | J1 | 40-44 | F |
| A3 | 225 | 253(=162+91) | 0 | 0 | 0 | 3 | J2 | 40-44 | F |
| A3 | 253 | 286(=225+61) | 0 | 0 | 0 | rest | J2 | 40-44 | F |
| A3 | 286 | 314 | 0 | 0 | 0 | rest | J3 | 40-44 | F |

For better understanding, here we show a complete data record of four sample subjects (A0,A1,A2,A3) determined to the infection analysis. All of them are recorded from T=0 (26.12.2020) until T=314 (4.11.2021).

A0 is not vaccinated and gets infected at the last day of the study. A1 has not been infected before, but gets infected (Day 140) and dies of covid (Day 150) before being vaccinated. A2 became first-dose vaccinated with Comirnaty on day 114, was infected between the first and second dose (day 142), got the second dose (day 220) and survives until the end of the study. A3 has been infected 20 days before beginning of the study, gets vaccinated by Janssen (Day 150) and is not infected until the end.

The input of `coxph` routine is displayed in Table S1. Note that there is typically more records then events as each follow-up covariate has to have its own interval.

**Table S2.** Details on analyses. *X*DeltaInf – a dummy equal to one if the VaccStatus value corresponds to vaccine *X* and the interval *T* 1 ⩾ Jul-01-2021, ^*^ – 61 days periods (otherwise 91 day periods for InfPrior).

| Analysis | Ages | Event | Covariates |
| --- | --- | --- | --- |
| Infections | all | Infected | $\text{InfPrior}$, $\text{VaccStatus}$, Sex, AgeGr |
| Reinfections | all | Infected | $\text{InfPrior}^*$, Sex, AgeGr |
| Hospitalizations | all | Hospitalized | $\text{VaccStatus}$, Sex, AgeGr |
| Deaths | all | DeadByCov | $\text{VaccStatus}$, Sex, AgeGr |
| Boosters | all | Infected | $\text{InfPrior}$, $\text{VaccStatus}$, Sex, AgeGr |
| Delta March | 70–79 | Infected | $\text{InfPrior}$, $\text{VaccStatus}$, Sex, AgeGr, $\text{ADeltaInf}$,<br>$\text{JDeltaInf}$, $\text{MDeltaInf}$, $\text{PDeltaInf}$ |
| Delta April | 55–69 | Infected | dtto. |
| Delta May | 30–54 | Infected | dtto. |

## Supplementary tables

**Table S3.** Vaccination timeline according to the age and professional prioritizations in the Czech Republic.

| Age category | Vaccination start | Professional and other categories | Vaccination start |
| --- | --- | --- | --- |
| 80+ | 2021-01-15 | critical infrastructure | 2020-12-27 |
| 70+ | 2021-03-01 | health-care workers | 2021-01-26 |
| 65+ | 2021-04-14 | pedagogical staff | 2021-02-27 |
| 60+ | 2021-04-23 | persons with chronic disease 1. priority | 2021-03-24 |
| 55+ | 2021-04-28 | persons with chronic disease 2. priority | 2021-04-12 |
| 50+ | 2021-05-05 | social workers | 2021-04-07 |
| 45+ | 2021-05-11 | academic staff | 2021-05-03 |
| 40+ | 2021-05-17 | foreigners | 2021-06-11 |
| 35+ | 2021-05-24 |  |  |
| 30+ | 2021-05-26 |  |  |
| 16+ | 2021-06-04 | boosters for prioritized | 2021-09-02 |
| 12+ | 2021-07-01 | boosters | 2021-09-20 |

**Table S4.**
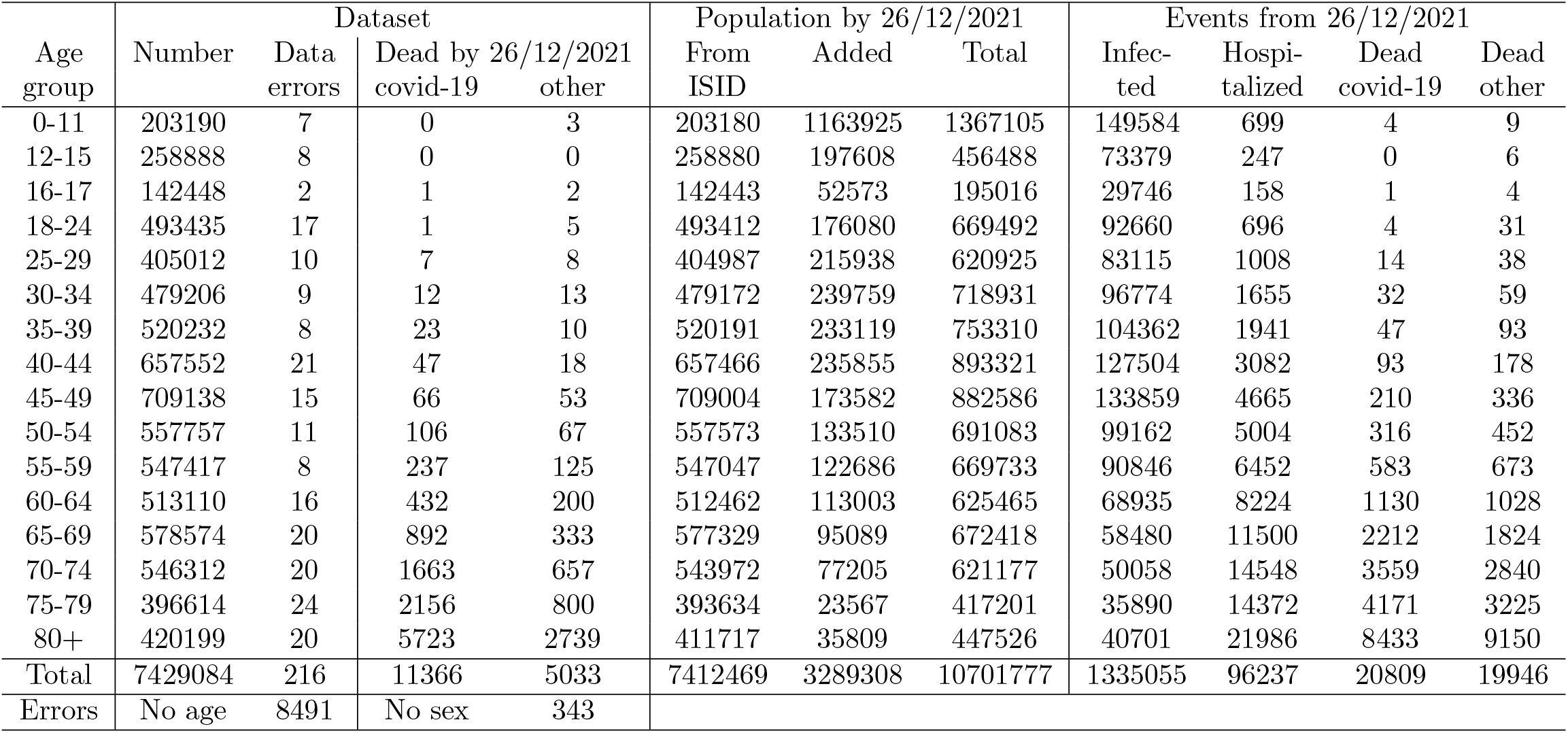
Descriptive statistics: population and epidemic characteristics in the Czech Republic; ISID = the Czech National Information System of Infectious Diseases.

**Table S5.** Descriptive statistics: vaccine distribution among different age groups until November 20, 2021.

| Age group | Completed vaccination |  |  |  |  | Booster doses |  |  |  |  |
| --- | --- | --- | --- | --- | --- | --- | --- | --- | --- | --- |
|  | Comirnaty | Spikevax | Vaxzevria | Janssen | Total | Comirnaty | Spikevax | Vaxzevria | Janssen | Total |
| 0-11 | 1435 | 27 | 2 | 4 | 1468 | 1 | 0 | 0 | 0 | 1 |
| 12-15 | 157523 | 2837 | 2 | 1 | 160363 | 7 | 1 | 0 | 0 | 8 |
| 16-17 | 105280 | 2249 | 39 | 306 | 107874 | 100 | 14 | 0 | 0 | 114 |
| 18-24 | 333076 | 22399 | 2446 | 39579 | 397500 | 4619 | 412 | 0 | 2 | 5033 |
| 25-29 | 261001 | 21720 | 3443 | 31346 | 317510 | 7999 | 695 | 2 | 5 | 8701 |
| 30-34 | 323970 | 24252 | 4864 | 32263 | 385349 | 9896 | 1039 | 4 | 5 | 10944 |
| 35-39 | 364023 | 27941 | 7219 | 33093 | 432276 | 14676 | 1617 | 2 | 10 | 16305 |
| 40-44 | 481994 | 36869 | 12470 | 37326 | 568659 | 23792 | 2580 | 13 | 19 | 26404 |
| 45-49 | 529313 | 40118 | 16480 | 39653 | 625564 | 30824 | 3384 | 7 | 17 | 34232 |
| 50-54 | 415101 | 34596 | 18013 | 31276 | 498986 | 28330 | 3255 | 9 | 19 | 31613 |
| 55-59 | 407622 | 34850 | 25000 | 29712 | 497184 | 32840 | 3742 | 10 | 24 | 36616 |
| 60-64 | 380452 | 34934 | 35010 | 27317 | 477713 | 31320 | 3758 | 5 | 24 | 35107 |
| 65-69 | 413874 | 44337 | 63135 | 27171 | 548517 | 33700 | 4876 | 11 | 31 | 38618 |
| 70-74 | 338152 | 56931 | 103284 | 19660 | 518027 | 110182 | 15891 | 35 | 56 | 126164 |
| 75-79 | 234084 | 45325 | 81112 | 11940 | 372461 | 125067 | 16403 | 24 | 62 | 141556 |
| 80+ | 264215 | 40220 | 64056 | 9414 | 377905 | 163649 | 18402 | 54 | 58 | 182163 |
| Total | 5011115 | 469605 | 436575 | 370061 | 6287356 | 617002 | 76069 | 176 | 332 | 693579 |

**Table S6.** Model outcomes for the effect of vaccination against infection, hospitalization and death. Hazard ratios together with 95% confidence intervals are provided. See the accompanying text for the meaning of model covariates.

| Model covariate | Infections | Hospitalizations | Deaths |
| --- | --- | --- | --- |
| AgeGr0-11 | 0.79 (0.78, 0.8) | 0.01 (0.01, 0.01) | 0 (0, 0) |
| AgeGr12-15 | 1.33 (1.32, 1.35) | 0.01 (0.01, 0.01) | 0 (0, 0) |
| AgeGr16-17 | 1.37 (1.35, 1.39) | 0.01 (0.01, 0.01) | 0 (0, 0) |
| AgeGr18-24 | 1.28 (1.26, 1.29) | 0.01 (0.01, 0.01) | 0 (0, 0) |
| AgeGr25-29 | 1.21 (1.19, 1.22) | 0.02 (0.02, 0.02) | 0 (0, 0) |
| AgeGr30-34 | 1.21 (1.2, 1.23) | 0.03 (0.03, 0.03) | 0 (0, 0) |
| AgeGr35-39 | 1.27 (1.25, 1.28) | 0.03 (0.03, 0.04) | 0 (0, 0) |
| AgeGr40-44 | 1.35 (1.33, 1.36) | 0.05 (0.05, 0.05) | 0 (0, 0) |
| AgeGr45-49 | 1.49 (1.48, 1.51) | 0.07 (0.07, 0.08) | 0.01 (0.01, 0.01) |
| AgeGr50-54 | 1.41 (1.4, 1.43) | 0.1 (0.1, 0.11) | 0.02 (0.01, 0.02) |
| AgeGr55-59 | 1.33 (1.32, 1.35) | 0.14 (0.14, 0.14) | 0.03 (0.03, 0.03) |
| AgeGr60-64 | 1.05 (1.04, 1.07) | 0.19 (0.19, 0.2) | 0.06 (0.06, 0.07) |
| AgeGr65-69 | 0.82 (0.81, 0.83) | 0.26 (0.25, 0.26) | 0.12 (0.11, 0.12) |
| AgeGr70-74 | 0.76 (0.75, 0.77) | 0.37 (0.36, 0.38) | 0.22 (0.21, 0.23) |
| AgeGr75-79 | 0.85 (0.83, 0.86) | 0.58 (0.57, 0.6) | 0.42 (0.4, 0.43) |
| SexZ | 1.06 (1.05, 1.06) | 0.69 (0.68, 0.69) | 0.47 (0.46, 0.49) |
| VaccStatusA_first1 | 0.64 (0.62, 0.67) | 0.42 (0.39, 0.45) | 0.36 (0.31, 0.4) |
| VaccStatusA_first2plus | 0.39 (0.34, 0.44) | 0.34 (0.27, 0.44) | 0.34 (0.22, 0.53) |
| VaccStatusA1 | 0.17 (0.15, 0.2) | 0.13 (0.09, 0.19) | 0.07 (0.02, 0.23) |
| VaccStatusA2 | 0.28 (0.27, 0.29) | 0.23 (0.2, 0.25) | 0.13 (0.1, 0.18) |
| VaccStatusA3 | 0.45 (0.44, 0.46) | 0.3 (0.28, 0.32) | 0.18 (0.15, 0.22) |
| VaccStatusA4 | 0.41 (0.3, 0.55) | 0.3 (0.14, 0.63) | 0.32 (0.08, 1.29) |
| VaccStatusJ1 | 0.32 (0.3, 0.34) | 0.32 (0.25, 0.4) | 0.32 (0.18, 0.58) |
| VaccStatusJ2 | 0.3 (0.29, 0.32) | 0.26 (0.22, 0.32) | 0.25 (0.15, 0.42) |
| VaccStatusJ3 | 0.33 (0.31, 0.35) | 0.33 (0.28, 0.38) | 0.32 (0.22, 0.47) |
| VaccStatusJ4 | 0.31 (0.23, 0.41) | 0.16 (0.06, 0.41) | 0.16 (0.02, 1.15) |
| VaccStatusM_first1 | 0.34 (0.32, 0.37) | 0.35 (0.31, 0.39) | 0.31 (0.26, 0.39) |
| VaccStatusM_first2plus | 0.23 (0.18, 0.3) | 0.21 (0.1, 0.43) | 0.24 (0.06, 0.96) |
| VaccStatusM1 | 0.1 (0.09, 0.11) | 0.06 (0.04, 0.08) | 0.04 (0.02, 0.09) |
| VaccStatusM2 | 0.15 (0.14, 0.15) | 0.1 (0.08, 0.13) | 0.04 (0.01, 0.1) |
| VaccStatusM3 | 0.22 (0.21, 0.23) | 0.14 (0.12, 0.16) | 0.09 (0.06, 0.14) |
| VaccStatusM4 | 0.35 (0.33, 0.37) | 0.19 (0.16, 0.22) | 0.12 (0.08, 0.18) |
| VaccStatusMboost | 0.07 (0.05, 0.09) | 0.02 (0.01, 0.05) | 0 (0, 0) |
| VaccStatusP_first1 | 0.52 (0.51, 0.53) | 0.39 (0.37, 0.41) | 0.38 (0.35, 0.41) |
| VaccStatusP_first2plus | 0.4 (0.37, 0.43) | 0.33 (0.25, 0.43) | 0.22 (0.11, 0.42) |
| VaccStatusP1 | 0.13 (0.13, 0.14) | 0.1 (0.09, 0.11) | 0.08 (0.07, 0.1) |
| VaccStatusP2 | 0.26 (0.25, 0.26) | 0.09 (0.08, 0.1) | 0.07 (0.06, 0.1) |
| VaccStatusP3 | 0.32 (0.32, 0.33) | 0.14 (0.13, 0.15) | 0.1 (0.08, 0.12) |
| VaccStatusP4 | 0.47 (0.46, 0.48) | 0.25 (0.24, 0.27) | 0.17 (0.14, 0.19) |
| VaccStatusPboost | 0.08 (0.08, 0.09) | 0.05 (0.04, 0.06) | 0.03 (0.02, 0.04) |
| InfPrior1 | 0 (0, 0) |  |  |
| InfPrior2 | 0.09 (0.09, 0.09) |  |  |
| InfPrior3 | 0.09 (0.09, 0.09) |  |  |
| InfPriorrest | 0.16 (0.16, 0.17) |  |  |

**Table S7.** Infection-acquired immunity against reinfection.

| Model covariate | Hazard ratio |
| --- | --- |
| AgeGr0-11 | 0.6 (0.6, 0.61) |
| AgeGr12-15 | 0.99 (0.98, 1) |
| AgeGr16-17 | 1.05 (1.03, 1.06) |
| AgeGr18-24 | 1.1 (1.09, 1.11) |
| AgeGr25-29 | 1.06 (1.05, 1.07) |
| AgeGr30-34 | 1.01 (1, 1.02) |
| AgeGr35-39 | 1.03 (1.02, 1.04) |
| AgeGr40-44 | 1.11 (1.1, 1.12) |
| AgeGr45-49 | 1.24 (1.23, 1.25) |
| AgeGr50-54 | 1.21 (1.2, 1.22) |
| AgeGr55-59 | 1.15 (1.14, 1.16) |
| AgeGr60-64 | 0.89 (0.88, 0.9) |
| AgeGr65-69 | 0.67 (0.66, 0.68) |
| AgeGr70-74 | 0.64 (0.63, 0.65) |
| AgeGr75-79 | 0.72 (0.71, 0.73) |
| InfPrior1 | 0 (0, 0) |
| InfPrior2 | 0.03 (0.03, 0.03) |
| InfPrior3 | 0.09 (0.09, 0.1) |
| InfPrior4 | 0.08 (0.08, 0.09) |
| InfPrior5 | 0.1 (0.09, 0.1) |
| InfPrior6 | 0.17 (0.16, 0.18) |
| InfPrior7 | 0.21 (0.2, 0.22) |
| InfPrior8 | 0.29 (0.27, 0.32) |
| InfPrior9 | 0.28 (0.22, 0.35) |
| InfPriorrest | 0.16 (0.12, 0.21) |
| SexZ | 1.06 (1.06, 1.06) |

**Table S8.** Observation numbers and average time intervals (in days) spent in individual periods used in the model for vaccination-induced immunity.

| Age cohort<br>Period | 12-24 years |  | 25-39 years |  | 40-65 years |  | 65+ years |  | Total |  |
| --- | --- | --- | --- | --- | --- | --- | --- | --- | --- | --- |
|  | number | time | number | time | number | time | number | time | number | time |
| A_first1 | 2574 | 57.5179 | 15927 | 58.2576 | 108526 | 59.0197 | 318607 | 59.9907 | 445636 | 59.678 |
| A_first2plus | 2062 | 23.2779 | 13306 | 24.4873 | 93899 | 22.4647 | 293687 | 23.7395 | 402956 | 23.4646 |
| A1 | 2486 | 60.5559 | 15519 | 60.7471 | 106945 | 60.8994 | 311509 | 60.9459 | 436461 | 60.9252 |
| A2 | 2438 | 55.7034 | 15331 | 56.2852 | 106417 | 54.4508 | 310785 | 57.8167 | 434973 | 56.9274 |
| A3 | 1870 | 34.3684 | 12119 | 34.3471 | 70806 | 30.8478 | 241225 | 27.8156 | 326022 | 28.7545 |
| A4 | 101 | 10.8416 | 568 | 15.0898 | 1613 | 13.9442 | 3783 | 11.0592 | 6065 | 12.2003 |
| J1 | 30377 | 44.4078 | 73480 | 43.6159 | 139716 | 48.277 | 63962 | 56.1901 | 307538 | 48.4269 |
| J2 | 17785 | 36.1091 | 42179 | 38.6196 | 95148 | 44.3658 | 54895 | 52.0908 | 210009 | 44.5316 |
| J3 | 4004 | 25.3469 | 11983 | 28.0255 | 42523 | 34.7839 | 37515 | 38.614 | 96025 | 35.0434 |
| J4 | 137 | 11.5401 | 483 | 6.33126 | 5468 | 4.65838 | 8928 | 5.12634 | 15016 | 5.05321 |
| M_first1 | 28947 | 31.0729 | 76818 | 32.6187 | 185331 | 34.057 | 189602 | 33.1329 | 480728 | 33.2826 |
| M_first2plus | 564 | 42.3777 | 1546 | 53.511 | 2538 | 60.0536 | 3155 | 90.7975 | 7804 | 69.9036 |
| M1 | 26472 | 55.5781 | 72370 | 59.0386 | 179160 | 59.9826 | 186121 | 60.5959 | 464145 | 59.8287 |
| M2 | 21440 | 46.9746 | 66917 | 50.6157 | 172514 | 56.3075 | 183564 | 59.4828 | 444436 | 56.3118 |
| M3 | 6545 | 27.5694 | 29717 | 31.7158 | 123056 | 30.2666 | 165658 | 47.4067 | 324977 | 39.0819 |
| M4 | 1902 | 26.0221 | 10606 | 25.2845 | 35976 | 26.1274 | 88390 | 23.8829 | 136874 | 24.6112 |
| Mboost | 213 | 10.9859 | 1732 | 10.4284 | 8903 | 10.6946 | 33080 | 10.1518 | 43928 | 10.2768 |
| P_first1 | 614063 | 31.038 | 971192 | 35.8558 | 2237875 | 37.806 | 1261365 | 33.1621 | 5086021 | 35.4599 |
| P_first2plus | 7903 | 38.5345 | 17016 | 51.1134 | 25837 | 53.3482 | 17484 | 81.8353 | 68248 | 58.3718 |
| P1 | 569712 | 57.705 | 922918 | 59.5366 | 2186557 | 60.383 | 1243178 | 60.6162 | 4923610 | 59.9708 |
| P2 | 483691 | 29.9072 | 867851 | 43.8328 | 2131571 | 55.5763 | 1225612 | 59.0829 | 4709451 | 51.6825 |
| P3 | 40873 | 36.0131 | 204188 | 36.0098 | 1532525 | 23.8681 | 1132070 | 44.6301 | 2909673 | 32.9687 |
| P4 | 15202 | 39.1274 | 78498 | 42.2502 | 253671 | 39.5429 | 535775 | 33.4437 | 883154 | 36.0763 |
| Pboost | 2426 | 15.7275 | 15054 | 16.9903 | 75979 | 15.512 | 275070 | 12.5485 | 368530 | 13.3619 |

## References

Abu-Raddad, LJ and Bertollini, R. (2021). Severity of SARS-CoV-2 reinfections as compared with primary infections. N Engl J Med,doi:10.1056/NEJMc2108120.

Baden, LR, Sahly, HM El, Essink, B, Kotloff, K, Frey, S and et al. (2021). Efficacy and safety of the mRNA-1273 SARS-CoV-2 vaccine. N Engl J Med 384, 403–416.

Bar-On, YM, Goldberg, Y, Mandel, M, Bodenheimer, O, Freedman, L, Kalkstein, N, Mizrahi, B, Alroy-Preis, S, Ash, N, Milo, R and others. (2021). Protection of BNT162b2 vaccine booster against Covid-19 in Israel. N Engl J Med 385, 1393–1400.

Brown, CM, Vostok, J, Johnson, H, Burns, M, Gharpure, R and et al. (2021). Outbreak of SARS-CoV-2 infections, including COVID-19 vaccine breakthrough infections, associated with large public gatherings - Barnstable County, Massachusetts, July 2021. MMWR Morb Mortal Wkly Rep 70, 1059–1062.

Cavanaugh, AM, Spicer, KB, Thoroughman, D, Glick, C and Winter, K. (2021). Reduced risk of reinfection with SARS-CoV-2 after COVID-19 vaccination — Kentucky, May–June 2021. MMWR Morb Mortal Wkly Rep 70, 1081–1083.

Chemaitelly, H, Tang, P, Hasan, MR, AlMukdad, S, Yassine, HM and et al. (2021). Waning of BNT162b2 vaccine protection against SARS-CoV-2 infection in Qatar. N Engl J Med,doi: 10.1056/NEJMoa2114114.

Cohn, BA, Circillo, PM, Murphy, CC, Krigbaum, NY and Wallace, AW. (2021). SARS-CoV-2 vaccine protection and deaths among US veterans during 2021. Science, doi: 10.1126/science.abm0620.

Hansen, CH, Michlmayr, D, Gubbels, SM, Mølbak, K and Ethelberg, S. (2021). Assessment of protection against reinfection with SARS-CoV-2 among 4 million PCR-tested individuals in Denmark in 2020: a population-level observational study. Lancet 397, 1204–1212.

Komenda, M, Bulhart, V, Karolyi, M, Jarkovský, J, Mužík, J and et al. (2020). Complex reporting of the COVID-19 epidemic in the Czech Republic: Use of an interactive web-based app in practice. J Med Internet Res 22, e19367.

Levine-Tiefenbrun, M, Yelin, I, Alapi, H, Katz, R, Herzel, E, Kuint, J, Chodick, G, Gazit, S, Patalon, T and Kishony, R. (2021). Viral loads of delta-variant SARS-CoV-2 breakthrough infections after vaccination and booster with BNT162b2. Nat Med, doi: 10.1038/s41591-021-01575-4.

Lopez Bernal, J. Andrews, N, Gower, C, Gallagher, E, Simmons, R, Thelwall, S, Stowe, J, Tessier, E, Groves, N, Dabrera, G, Myers, R, Campbell, CNJ, Amirthalingam, G, Edmunds, M, Zambon, M, Brown, KE, Hopkins, S, Chand, M and others. (2021). Effectiveness of Covid-19 vaccines against the B.1.617.2 (delta) variant. New Engl J Med 385(7), 585–594.

Ministry of Health of the Czech Republic. (2020). COVID-19: an overview of the actual situation in the Czech Republic (in Czech). onemocneni-aktualne.mzcr.cz/covid-19. Accessed: 2021-11-20.

Polack, FP, Thomas, SJ, Kitchin, N, Absalon, J, Gurtman, A and et al. (2020). Safety and efficacy of the BNT162b2 mRNA Covid-19 vaccine. N Engl J Med 383, 2603–2615.

Sadoff, J, Gray, G, Vandebosch, A, Cárdenas, V, Shukarev, G and et al. (2021). Safety and efficacy of single-dose Ad26.COV2.S vaccine against Covid-19. N Engl J Med 384, 2187–2201.

Tang, P, Hasan, MR, Chemaitelly, H, Yassine, HM, Benslimane, FM, Al Khatib, HA, AlMukdad, S, Coyle, P, Ayoub, HH, Al Kanaani, Z, Al Kuwari, E, Jeremijenko, A, Kaleeckal, AH, Latif, AN, Shaik, RM, RHF, Abdul, Nasrallah, GK, Al Kuwari, MG, Al Romaihi, HE, Butt, AA, Al-Thani, MH, Al Khal, A, Bertollini, R and others. (2021). BNT162b2 and mRNA-1273 COVID-19 vaccine effectiveness against the SARS-CoV-2 delta variant in Qatar. Nat Med,doi: 10.1038/s41591-021-01583-4.

Tartof, SY, Slezak, JM, Fischer, H, Hong, V, Ackerson, BK, Ranasinghe, ON, Frankland, TB, Ogun, OA, Zamparo, JM, Gray, S and others. (2021). Effectiveness of mRNA BNT162b2 COVID-19 vaccine up to 6 months in a large integrated health system in the USA: a retrospective cohort study. Lancet 398, 1407–1416.

Townsend, JP, Hassler, HB, Wang, Z, Miura, S, Singh, J, Kumar, S and et al. (2021). The durability of immunity against reinfection by SARS-CoV-2: a comparative evolutionary study. Lancet Microbe,doi: 10.1016/S2666-5247(21)00219-6.

Voysey, M, Clemens, SA Costa, Madhi, SA, Weckx, LY, Folegatti, PM, Aley, PK and et al. (2021). Safety and efficacy of the ChAdOx1 nCoV-19 vaccine (AZD1222) against SARS-CoV- 2: an interim analysis of four randomised controlled trials in Brazil, South Africa, and the UK. Lancet 397, 99–111.

